# INTERCEPT pathogen reduction of platelet concentrates induces trans-arachidonic acids and affects eicosanoid formation

**DOI:** 10.1101/2022.04.06.22273484

**Authors:** Gerda C. Leitner, Gerhard Hagn, Laura Niederstätter, Andrea Bileck, Kerstin Plessl-Walder, Michaela Horvath, Vera Kolovratova, Andreas Tanzmann, Alexander Tolios, Werner Rabitsch, Philipp Wohlfarth, Christopher Gerner

## Abstract

Gamma-irradiation of blood products is mandatory to avoid graft versus host disease in patients with immunosuppressed clinical conditions. Pathogen inactivation techniques were implemented to optimize safe blood component supply. The INTERCEPT treatment uses amotosalen together with UVA irradiation. The functional and molecular implications of these essential treatments have not yet been systematically assessed. The irradiation-induced inactivation of nucleic acids may actually be accompanied with modifications of chemically reactive polyunsaturated fatty acids, known to be important mediators of platelet functions. Thus, here we investigated eicosanoids and related fatty acids released upon treatment and during platelet storage for 7 days, complemented by the analysis of functional and metabolic consequences of these treatments. In contrast to gamma-irradiation, here we demonstrate that UVA treatment attenuated the formation of ALOX12-products such as 12-HETE and 12-HEPE but induced the formation of trans-arachidonic acids in addition to 11-HETE and HpODEs. Metabolic and functional issues like glucose consumption, lactate formation, platelet aggregation and clot firmness hardly differed between the two treatment groups. *In vitro* synthesis of trans-arachidonic acids (trans-AA) out of arachidonic acid in the presence of β-mercaptoethanol suggested that thiol radicals formed by UVA treatment are responsible for the INTERCEPT-specific effects observed in platelet concentrates. It is plausible to assume that trans-AA and other UVA-induced molecules may have specific biological effects on the recipients, which need to be addressed in future studies.

**Key points:**

- A previously unrecognized radical mechanisms for the generation of trans-fatty acids by UVA was identified
- Irradiation with UVA was found to immediately affect the generation of polyunsaturated fatty acid oxidation products

## Introduction

One of the key elements of blood transfusion is the availability of safe blood and blood products. In addition to the careful and sophisticated donor selection the application of state-of-the-art blood processing procedures and testing is mandatory.

Transfusion associated graft versus host disease (TA-GvHD) is a very rare but usually fatal complication following transfusion of lymphocyte-containing blood components in immunosuppressed patients.^1-3^ But also non-immunosuppressed patients could experience this problem, particularly if the transfused blood components were donated from an HLA haploidentical unrelated donor or family member.^4-8^ Since the early 1980ties the irradiation of blood components with at least 25 Gy is strongly recommended for patients at risk to develop TA-GvHD.^9^ Gamma-irradiation inactivates residual T-cells in blood components, the source cells for TA-GvHD. However, gamma irradiation (yIRR) may induce metabolic injuries and reduces shelf life of blood components.^10^

Over the last decades, big efforts were made to further increase blood safety and reduce the risk for transfusion transmitted infections (TTIs). The most recent innovation in this field was the development of pathogen reduction (PR) methods and inactivation (PI) techniques for platelet concentrates (PCs) and plasma. These techniques are effective against proliferating nucleated cells of pathogens, thus also effective against TA-GvHD inducing T-cells. Consequently, irradiation of blood components in patients at risk can be omitted. Several studies were carried out to prove the hemostatic efficacy of PR or PI platelets.^11-13^ However, while some studies reported that post-transfusion increment of platelet counts was significantly lower after transfusion of PR and PI platelets, this was no consistent observation.^14,15^ Anyhow, concerns about function of these pre-treated platelets arose.

Here we investigated the *in vitro* properties of either yIRR- or INTERCEPT-treated PCs by measuring metabolic changes and the formation of eicosanoids.

Polyunsaturated fatty acids are highly plausible candidate molecules present in platelets which may show some chemical reactivity upon irradiation.^16^ Arachidonic acid is a polyunsaturated fatty acid abundantly esterified in membrane lipids such as phosphatidylcholines. Oxidation of arachidonic acid may occur in an enzymatic or non-enzymatic fashion, eventually forming so called eicosanoids, which are known to be of high functional relevance for platelets.^17^ Thromboxane A2 is known as the main prostanoid promoting platelet aggregation, which is modulated by various prostaglandins and hydroxyeicosatetraenoic acids (HETEs).^18^ The eicosanoid background may thus significantly affect the coagulation properties of the transfused platelet concentrates.^19^ Thus, here we investigated the release of eicosanoids and related molecules in PCs before and after yIRR or INTERCEPT treatment, using an untargeted high-resolution mass spectrometry-based method as employed by us previously.^20^

## Methods

### Study design

This study was an *in vitro* investigation of PCs conducted at the Medical University of Vienna after prior approval of the local ethics committee. Fifteen double-unit PCs were collected from 15 healthy volunteers of our multicomponent donor pool. All donors gave written informed consent to participate in this study. All PCs were divided into two single units so that the requirements for PI of a single unit was fulfilled. The second smaller unit was transferred into an oxygen permeable storage bag from TerumoBCT (Denver, CO, USA). One unit was pathogen inactivated and one unit was irradiated with 30 Gy. PCs were stored at 22 °C (–2 °C) under continuous agitation on an agitator (Helmer Platelet Incubator, Noblesville, IN, USA) for 7 days. Samples were drawn under aseptic conditions after production (day 0, before dividing into two parts), after either yIRR (group irradiated) or PI treatment (group intercept) (day 1), further on days 5 and 7 (end of shelf life). All units were tested on platelet counts, residual white blood cells (rWBC), pH, lactate, lactatdehydrogenase (LDH), glucose and thromboelastometry (TEM) measurements. TEM was performed to work out if both manipulations have an impact on hemostatic function of fresh and stored collected platelets or not. Sterility testing was done only on day 7. Additionally, all units were tested on the aggregation response to collagen and the thrombin receptor agonist peptide-6 (TRAP-6). PI treatment was performed with the Intercept™ Blood System/Cerus Europe B.V. All PCs were visually inspected for swirling and aggregates.

### Donors

All donors (11 males, 3 females) met the national and international requirements for healthy volunteers (Austrian law, EU guidelines), median age was 39 years (range 22-65). Double adult dose PCs were collected by the TrimaAccel collection system (version 6.0; TerumoBCT, Denver, CO, USA).

### Laboratory analyzes

Platelet (Plt) counts were measured with a hematology analyzer (Cell Dyn Ruby, Abbott, South Pasadena, CA, USA). pH was measured with a biochemistry analyzer at 22 °C (ABL 80 FLEX Analysator, Drott, Medizintechnik GmbH, Austria); lactate dehydrogenase (LDH) was analyzed with a Beckmann Coulter AU 5800 (Bremerhaven, Germany) and glucose and lactate with an OLYMPUS AU 5430 (Beckmann Coulter, Danvers, MA, USA).

### Platelet function and activation by Light transmission aggregometry (LTA)

Platelet function and activation by LTA was performed using a PAP 8E (MoeLab, Langenfeld, Germany), as previously described.^21,22^ In short, PCs were diluted to a final concentration of 250 ×10^9^/L with solvent-detergent plasma (Octaplas; blood group AB; Octapharma, Vienna, Austria). Aggregation response to collagen (final concentration 190 µg/mL; BioData Corporation, Horsham, PA, USA) or TRAP-6 (final concentration of 25 µmol; Bachem, Bubendorf, Switzerland) was continuously recorded for 5 min. The maximal aggregation (MA) was used for further statistics.

### Thromboelastometry

The ROTEM® (Pentapharm GmbH, Munich) is a point-of-care analyzer which uses normally citrated whole blood samples for the analyzation of the viscoelastic properties of the sample as it clots. Here PC samples were treated analog to LTA measurements to mimic whole blood. In our department only the NATEM test was performed. In this test, clot formation is initiated through activation of coagulation by contact. The clot formation time (CFT) and the maximal clot firmness (MCF) were regarded as the primary variables of interest, as both are mainly influenced by the number and the function of platelets.^23,24^ The CFT is defined as the time from clotting start until the clot firmness reaches the 20 mm mark. The maximal clot firmness (MCF) is the greatest vertical amplitude of the typical trace and reflects the absolute strength of the fibrin and platelet clot.^25^

### Residual leukocytes and sterility tests

Residual leukocytes were assessed only on the first day of storage by flow cytometry using the ‘leukocount’ kit (Becton Dickenson, San Jose, CA, USA). Sterility tests were performed by an accredited institute according to the requirements of Ph.Eur. 2.

### Pathogen inactivation of platelets

Our institution produces solely single-donor platelets by apheresis. We have been using the Intercept™ system for PI of PCs since March 2012. This system has been shown to effectively inactivate contamination of PCs with a broad spectrum of viruses, bacteria, and parasites as well as donor leukocytes. Our national authorities allow a 7-day storage for 100% PI PCs, which has therefore been implemented as maximum storage time at our institution. The Intercept™ system uses amotosalen HCl (a photoactive compound) and long-wavelength ultraviolet (UVA) illumination for *ex vivo* PI of PCs. Residual amotosalen and free photoproducts are reduced to low levels by exposure to a compound adsorption device (CAD) before transfer of the treated platelets to a storage container for release.

### Irradiation of platelets

The 30 gy irradiation of the second unit of PCs was done with an IBL 437 C (cis bio international, France). This device uses a Caesium 137 source.

### Sample preparation for eicosanoid analysis

1 mL platelet concentrate was transferred into a 15 mL falcon tube for the two different treatment types. 4 mL ice cold ethanol (EtOH) were added, containing 10-100 nM of each internal standard (12S-,15S-HETE-d8, 5-Oxo-ETE-d7, 11,12-DiHETrE-d11, PGE2-d4, 20-HETE-d6; Cayman Chemical, Tallinn, Estonia) for protein precipitation over night at -20 °C. Samples were then centrifuged for 30 min at 4536 g and 4 °C, protein pellets discarded and EtOH evaporated *via* vacuum centrifugation at 37 °C until the original sample volume was restored. Solid-phase extraction (SPE) was performed with 30 mg/mL StrataX SPE columns (Phenomenex, Torrance, CA, USA). Columns were washed with 2 mL absolute Methanol (abs. MeOH), equilibrated with 2 mL MS grade H_2_O before sample loading with Pasteur pipettes. Samples were washed with 5 mL MS grade H_2_O before eluting the eicosanoids into 1.5 mL glass vials with 500 µL abs. MeOH containing 2% formic acid (FA). The eluates were stored at -80 °C until further analysis. Before measurement, the organic phase was evaporated at room temperature using a steam of nitrogen and reconstituted in 150 µL reconstitution buffer (H_2_O/acetonitrile (ACN)/MeOH + 0.2% FA - 65:31.5:3.5), containing another set of 10-100 nM internal standards (5S-HETE-d8, 14,15-DiHETrE-d11, 8-iso-PGF2a-d4; Cayman Chemical, Tallinn, Estonia).

### Synthesis of trans-arachidonic acid

Trans-arachidonic acid was synthesised *via* isomerization of all cis-arachidonic acid according to Ferreri et al.,^26^ applying a slightly adapted protocol. In brief, a solution of 1.5 mM arachidonic acid in isopropanol containing 10 mM β-mercaptoethanol was placed in an iOS quartz cuvette and bubbled with a gentle stream of nitrogen for 75min, while exposing to 364 nm wavelength (UVP UVGL-58, Analytik Jena, Germany). Afterwards, isopropanol and β-mercaptoethanol were evaporated using nitrogen and the sample reconstituted in 150 µL reconstitution buffer containing internal standards as described above.

### LC-MS/MS analysis of eicosanoids

Separation of the analytes was achieved with an UHPLC system (Thermo Scientific™ Vanquish, Austria) equipped with a reversed phase C18 column (Kinetex® 2.6 µm XB-C18 100 Å, LC Column 150 × 2.1 mm, Torrance, CA, USA). The flow rate was set to 200 µL min^-1^, the HPLC oven temperature to 40 °C, the auto sampler to 4 °C and the injection volume was 20 µL. A gradient method with a total run time of 20 min was applied using eluent A (H_2_O + 0.1% FA) and eluent B (90% ACN + 10% MeOH + 0.1% FA). Starting condition of 35% B was kept constant for 1 minute, increasing linear up to 90% B within 9 minutes and then further increased up to 99% B within 0.5 minutes, where it was kept constant for 5 minutes, restoring initial conditions within 0.5 minutes.

MS analysis was performed using a high-resolution quadrupole orbitrap mass spectrometer (Thermo Scientific™ QExactive™ HF hybrid quadrupole-orbitrap mass spectrometer) equipped with a HESI source run in negative ionization mode. Data was recorded in full scan mode operating in the mass range *m/z* 250-700 at 60’000 resolution (*m/z* 200). A Top 2 method was chosen for MS/MS fragmentation in data dependent acquisition (DDA) mode and fragmentation was achieved using an HCD with a normalized collision energy of 24. Spray voltage was set to 2.2 kV, capillary temperature to 253°C and sheath gas and auxiliary gas to 46 and 10 arbitrary units, respectively.

### LC-MS/MS data processing

Identification of eicosanoids was performed manually using Xcalibur Qual Browser (Version 4.1.31.9; Thermo Fisher Scientific, Austria) by using the exact mass, retention time and MS/MS fragmentation pattern which where manually compared to the reference spectra on LIPIDMAPS depository library from July 2020.^27^ This putatively identified molecules are marked with its retention time after either the exact mass or its MS2 based proposed identification. External standards were then used for verification of the putatively identified fatty acids. In case of the presently described HpODEs, monoisotopic exact mass, isotopic pattern and fragmentation spectra clearly identified the molecular lipid but did not allow us to define the molecular structure. However, a comparison with commercial standards ruled out 9-HpODE and 13-HpODE, respectively. A lack of further commercially available standards did not yet allow us further clarification. For relative quantification the software TraceFinder (Version 4.1; Thermo Fisher Scientific, Austria) was applied, allowing a mass deviation of 5 ppm. For visualization and statistical evaluation, the software GraphPad Prism (Version 6.07) and Origin Pro (Version 2019b) was used.

### Statistics

Results are shown as mean and standard deviation (SD) or median and range for descriptive purposes in the text. All statistical comparisons were made by non-parametric tests due to the non-normal distribution of the data. Differences between the two groups (irradiated and pathogen inactivated PCs) and changes during storage were analyzed by Man Whitney U Test. For evaluation of metabolic changes within the groups over time the Wilcoxon paired signed rank test for dependent samples was applied. A two-sided p value <0.05 was considered statistically significant.

## Results

### Platelet metabolism and function are hardly affected by irradiation and INTERCEPT treatment

Glucose consumption may serve as an indicator for platelet metabolism and was evidenced by a steady decrease of glucose concentration accompanied by the formation of lactate during storage of platelet concentrates (Figure 1A, 1B). A comparison between irradiation and pathogen inactivation pointed to a slightly decreased formation of lactate at day 7 upon INTERCEPT treatment. Remarkably, clot formation time and clot firmness as well as the TRAP-induced maximal aggregation did not differ significantly between these two groups. However, collagen- and TRAP-6-induced aggregation decreased significantly over time (p < 0.05), whereas clot formation time and clot firmness remained stable upon the storage time (Table 1a, Figure 1 C, D).

**Table 1a:** Rotem data of irradiated and pathogen inactivated platelet units. (CFT, clot formation time (seconds); MCF, maximal clot firmness (millimeter); SD, standard deviation)

|  | irradiated |  | pathogen inactivated |  |
| --- | --- | --- | --- | --- |
|  | CFT sec<br>(mean/SD) | MCF mm<br>(mean/SD) | CFT sec<br>(mean/SD) | MCF mm<br>(mean/SD) |
| Day 1 | 68 (3) | 152 (22) | 68 (3) | 152 (22) |
| Day 2 | 68 (3) | 162 (33) | 66 (1) | 156 (29) |
| Day 5 | 68 (3) | 152 (40) | 67 (3) | 142 (21) |
| Day 7 | 67 (3) | 157 (23) | 65 (2) | 173 (27) |

**Table 1b:** Aggregometry results after collagen and TRAP-6 enhancement. (TRAP-6, thrombin receptor agonist peptide-6; MA, maximal aggregation; SD, standard deviation)

|  | irradiated |  | pathogen inactivated |  |
| --- | --- | --- | --- | --- |
|  | Collagen MA<br>(mean/SD) | TRAP-6 MA<br>(mean/SD) | Collagen MA<br>(mean/SD) | TRAP-6 MA<br>(mean/SD) |
| Day 1 | 88 (15) | 91 (18) | 88 (15) | 91 (18) |
| Day 2 | 81 (7) | 81 (6) | 81 (10) | 69 (17) |
| Day 5 | 74 (10) | 65 (15) | 78 (11) | 65 (24) |
| Day 7 | 68 (12) | 62 (12) | 69 (11) | 62 (14) |

**Figure 1:**
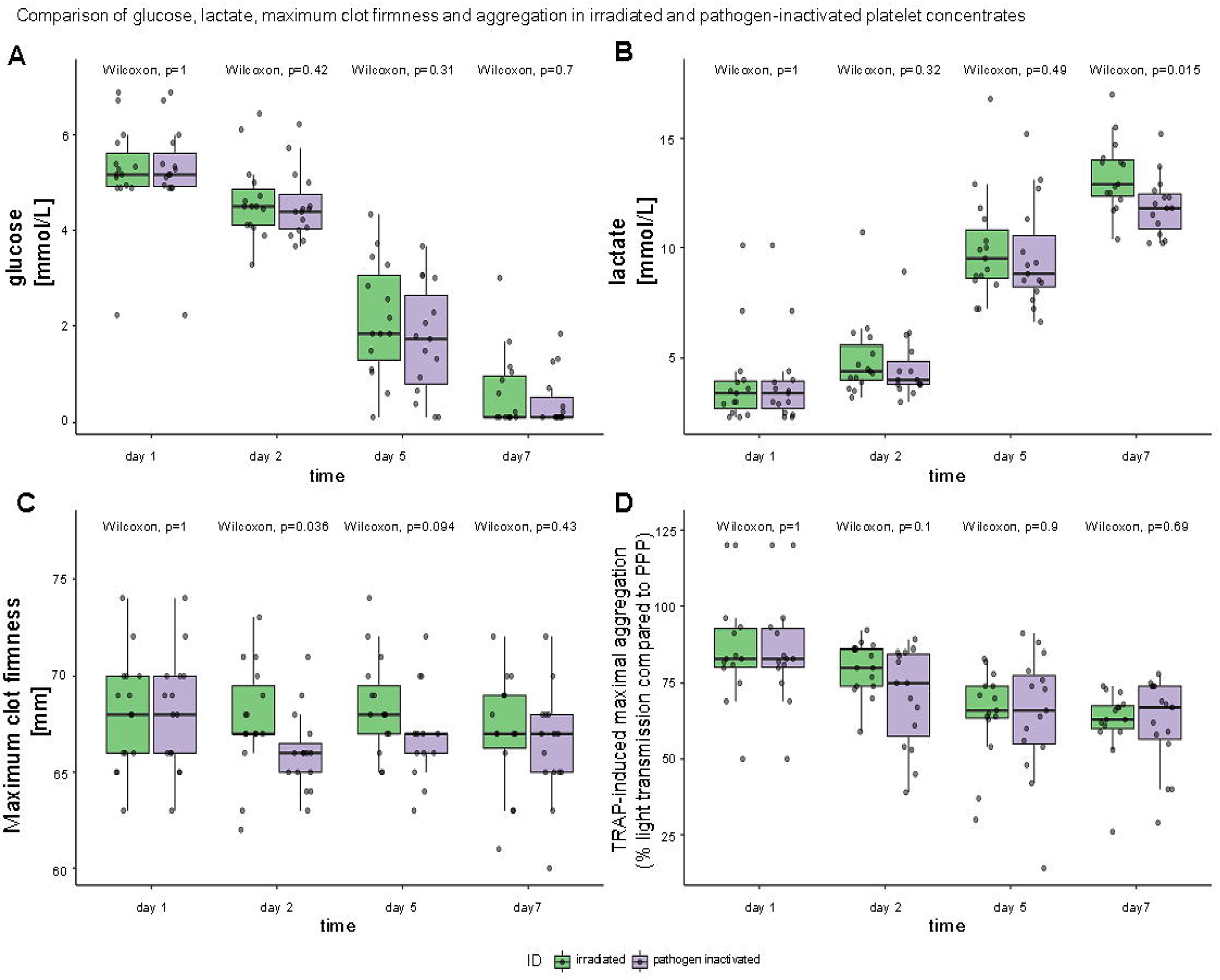
Comparison of **A**, glucose; **B**, lactate; **C**, maximum of clot firmness and **D**, aggregation of irradiated and pathogen-inactivated platelet concentrates.

### INTERCEPT treatment for pathogen reduction induces the specific formation of trans-arachidonic acid

Platelets are known to constantly release arachidonic acid, together with several eicosanoids such as 12S-HETE.^28^ Indeed, a steady release of AA and the 12LOX-products 12S-HETE and 12-HEPE was also observed by us during storage (Figure 2A). Remarkably, the 12LOX-products hydroperoxyoctadecadienoic acids (HpODEs), formed from linoleic acid rather than AA, were found apparently decreased upon UVA treatment, but increased upon y-irradiation and during storage (Fugure 2A, Suppl. Figure 1). Consistently, at day 5 and 7, the relative concentration of these molecules was significantly higher in gamma-irradiated platelet concentrates when compared to UVA-treatment (Figure 2A).

**Figure 2:**
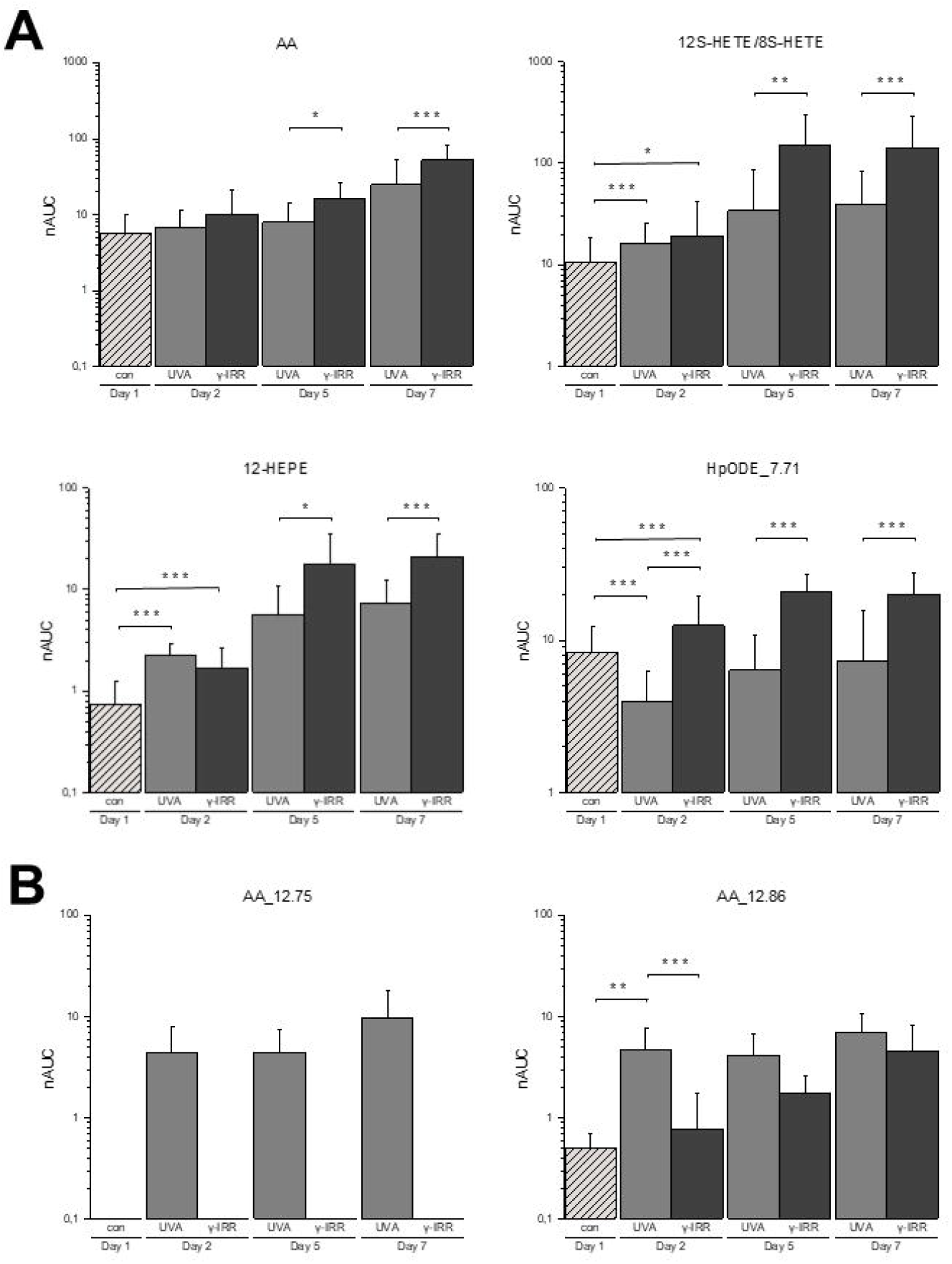
Time-dependent concentration changes of arachidonic acid (AA), 12S-HETE, 12-HEPE and a HpODE with an observed retention time of 7,71 minutes **(A)**. Transarachidonic acids with the retention times of 12,75 minutes and 12,86 minutes were specifically induced with UVA treatment of platelet concentrates **(B)**.

On the other hand, UVA-treated samples, but not gamma-irradiated samples, showed the specific formation of trans-arachidonic acids, currently distinguished by different retention times as observed by chromatography (Figure 2B, Suppl. Figure 1). The formation of trans-arachidonic acids was reported to occur either by the action of nitric oxide radicals *in vivo*^29^ or by thiol radicals formed using radical reaction initiators such as azide radical.^26^ Thus we wondered whether UV light might be capable of initiating the formation of similar sulfhydryl radicals in the absence of amotosalen or other potential radical reaction initiators. Under a constant flow of nitrogen gas in order to remove any solubilized oxygen, pure arachidonic acid was irradiated with the same dose of UVA as used for INTERCEPT treatment in the presence of β-mercaptoethanol as described in Materials. Remarkably, the same trans-AA molecules were indeed formed in this cell-free experiment as observed by INTERCEPT treatment of platelet concentrates (Figure 3).

**Figure 3:**
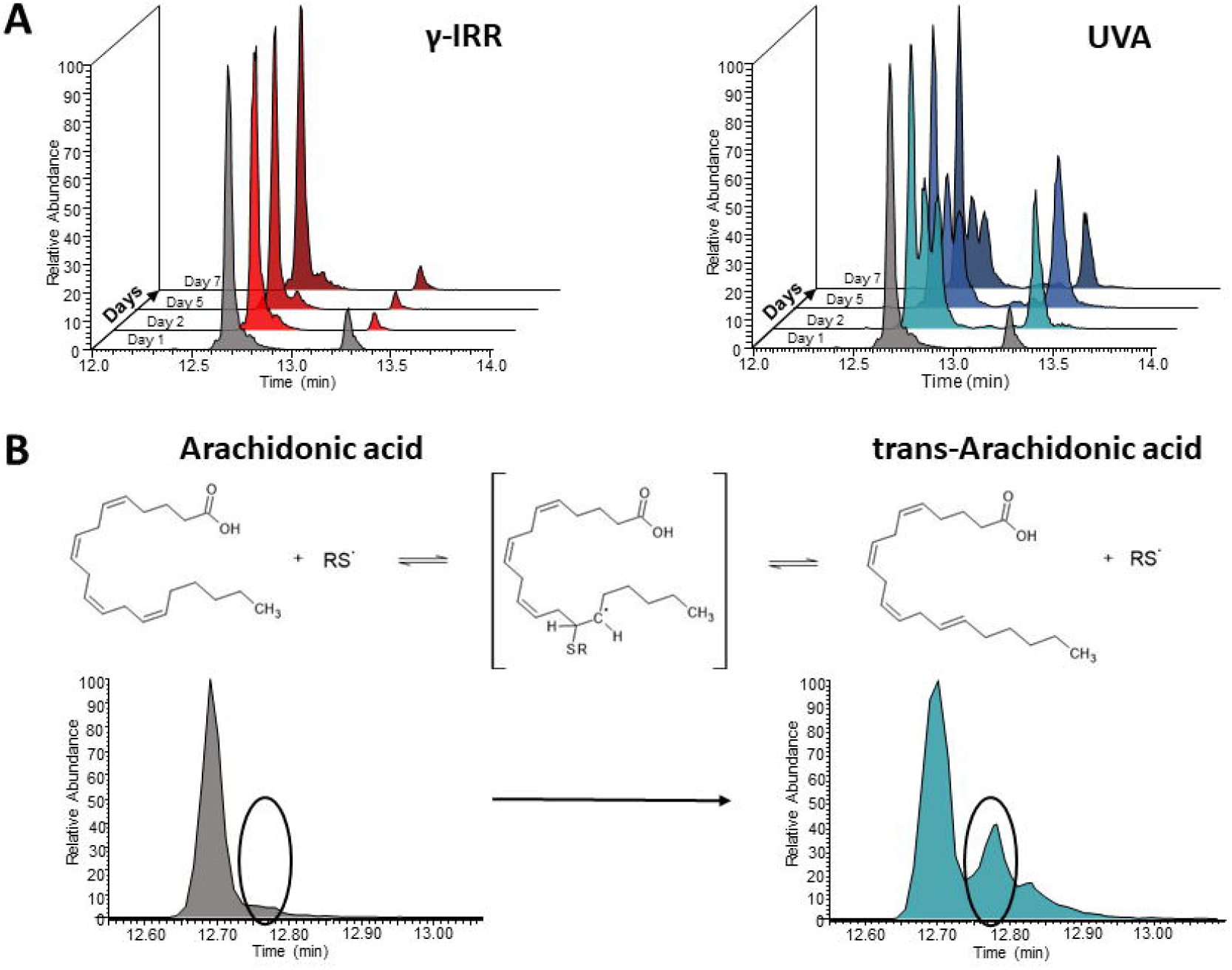
**A**, UVA treatment, but not γIRR, is accompanied with the formation of trans-arachidonic acid. **B**, the formation of trans-arachidonic acid was also obtained by UVA treatment of arachidonic acid with a sulfhydryl-donor (beta mercaptoethanol).

### INTERCEPT treatment also promotes the formation of hydroxyeicosatetraenoic acids (HETEs)

In addition to the trans-arachidonic acids, further eicosanoids were found altered upon platelet treatment and storage. 11S-HETE and 15S-HETE were positively identified in all samples, but significantly up-regulated upon UVA treatment (Figure 4A). In contrast, 16-HETE and 18-HETE were not identified before irradiation, but were found exclusively at day 2 in all UVA-treated, but not before treatment nor in γ-IRR-treated samples (Figure 4B). However, at later time points, also these eicosanoids were detected in γ-IRR-treated samples. Remarkably, γ-IRR-treatment did not directly induce the formation of any eicosanoid or eicosanoid-like features, as evidenced upon comparison of the respective analysis results before and after irradiation (Figure 4C).

**Figure 4:**
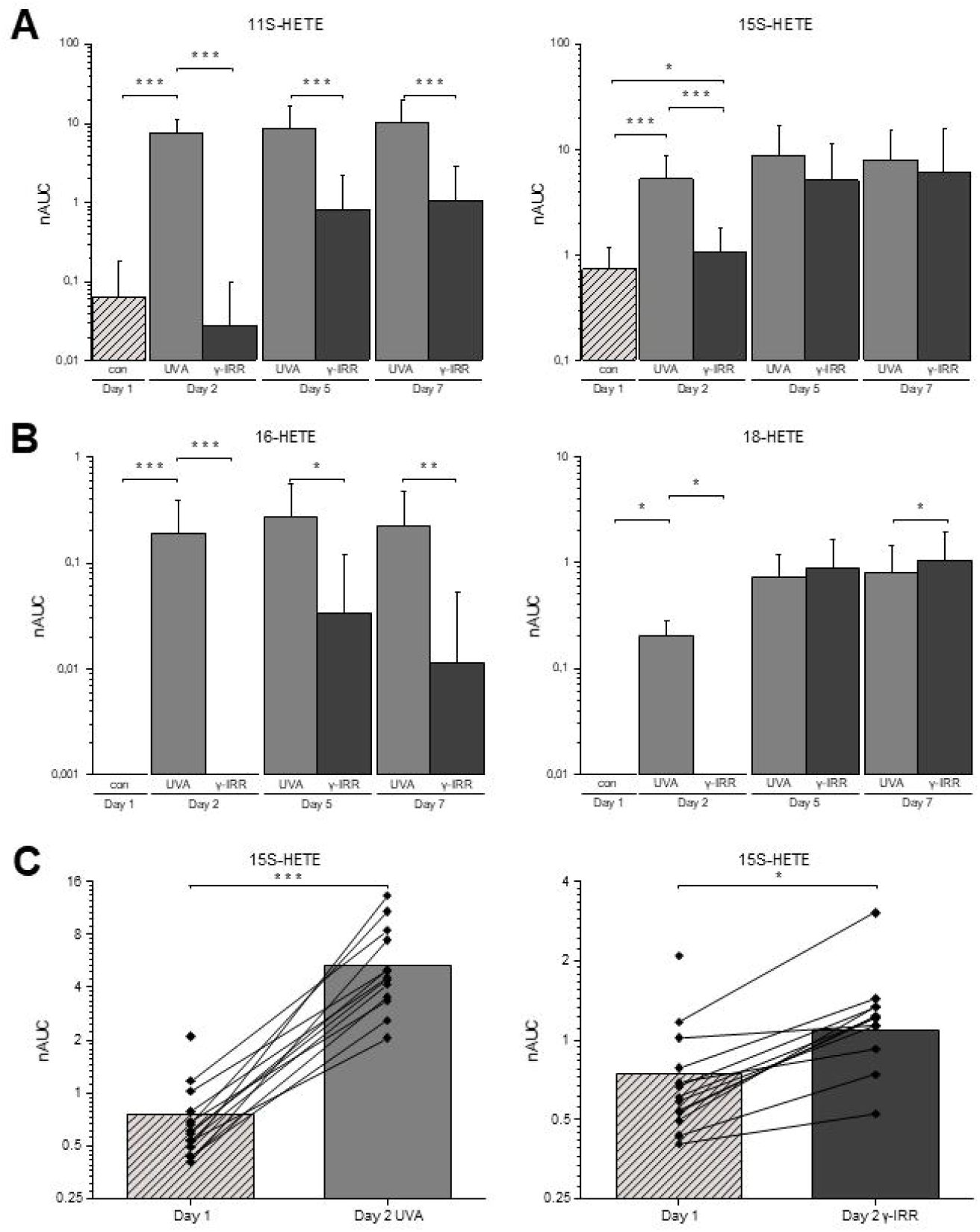
UVA treatment immediately increased levels of 11S-HETE and 15S-HETE **(A)**, and immediately induced the formation of 16-HETE and 18-HETE **(B)**. Note that these molecules were also observed upon γIRR, albeit at later time points. The relative increase was uniformly observed in all donors **(C)**.

## Discussion

As long as no stable *in vitro* model system for megakaryocyte culture and platelet formation is presented, patients with urgent demands for platelet concentrates will have to be treated with samples derived from individual donors, thus subject to substantial inter-individual variations. All parameters recorded in the present study with regard to platelet properties showed high degrees of inter-individual variations, which may indeed relate to variations in the clinical outcome of platelet treatment. Importantly, platelet concentrates have to be pretreated to avoid graft versus host disease for patients with severely immunosuppressed conditions (*i*.*e* after PBSC or bone marrow transplantation, newborns with SCID). The method of choice to inactivate T-cells is 25-30 Gy gamma irradiation (yIRR) of the products.^9^ Secondly, pathogen inactivation (PI; INTERCEPT) and pathogen reduction (PR) methods were implemented to minimize the risk of TTIs. These methods affect and inactivate nucleated cells and thus are also effective in inactivating the candidate cells for GvHD. As a consequence, in case of PI, yIRR can be omitted. However, these treatments may induce further variations which have not yet been investigated in molecular detail as presented here.

Eicosanoids are a group of lipid hormones formed by the oxidation of polyunsaturated fatty acids. Platelets are known producers of these highly bioactive molecules,^28^ motivating us to focus on this class of biomolecules. Importantly, eicosanoids are potent modulators of immune functions^30^ with direct effects on various leukocyte functions.^20^ The association with innate immune functions and various diseases including cancer^31,32^ makes these molecules potentially relevant for chronic diseases.

Here we demonstrate that INTERCEPT treatment directly causes the formation of trans-arachidonic acids (trans-AAs). Trans-AAs have been described to be formed by the action of nitric oxide radicals *in vivo*^*29*^ or catalyzed by sulfhydryl-groups in the presence of azide radical *in vitro*.^26^ Thus, here we wondered whether UVA treatment might replace azide radical and exposed pure arachidonic acid in the presence of β-mercaptoethanol to UVA light. Following this protocol, we were able to reproduce the trans-arachidonic acid pattern observed upon INTERCEPT treatment of platelet concentrates (Figure 3). Thus, here we suggest that UVA light in the presence of polyunsaturated fatty acids and sulfhydryl groups might induce the formation of sulfhydryl radicals which may cause the formation of trans-arachidonic acid.

In addition to trans-AAs, we also observed the increased formation of the COX1-product 11-HETE, the 15-LOX-product 15-HETE and the P450 products 16-HETE and 18-HETE upon UVA-treatment (Figure 4). This is in stark contrast to the formation of ALOX12-products 12S-HETE and 12-HEPE, which were significantly higher in gamma-irradiated samples (Figure 2). These observations point to a UVA-induced inhibition of ALOX12 activity as already described in case of keratinocytes exposed to sun light.^33^ Consequently, we suggested that the molecules apparently induced upon gamma-irradiation (Figure 2A) may rather result from the unaltered background enzymatic activity in course of platelet storage. Actually phospholipase A2 activity is known to cause constant release of PUFAs during platelet storage.^34^ These PUFAs are then subject to a broad variety of enzymatic or non-enzymatic oxidation processes, resulting in the formation of the oxylipins with remarkable bioactivities. The consumption of the precursor molecule arachidonic acid to form trans-AA upon UVA treatment in contrast to γ-irradiation may cause the relatively lower levels of these molecules in UVA-treated samples as displayed in Figure 2, which may superficially but wrongly suggest that γ-irradiation was promoting the formation of the ALOX12-products 12S-HETE, 12-HEPE and HpODE.

It is remarkable that on the other hand, P450 monooxygenase products 16-HETE and 18-HETE were specifically formed upon UVA treatment (Figure 4), pointing to increased enzymatic activity. An induction of P450 1A1 and P450 1B1 upon UV-exposure of human has been described^35^ and may be related to the present observation.

In summary, the present data indicate that UVA-treatment causes the formation of trans-arachidonic acid, promotes the formation of P450 products and inhibits the formation of ALOX12-products. We have not found evidence for a direct effect of gamma irradiation on PUFA processing and eicosanoid formation. Thus, the two treatment options of platelet concentrates are associated with marked differences in eicosanoid formation with potential clinical implications for the recipient which need to be addressed in future studies.

## Data Availability

All data produced in the present study are available upon reasonable request to the authors

## Abbreviations

AA: arachidonic acid
CAD: compound adsorption device
CFT: clot formation time
EtOH: ethanol
FA: formic acid
HETEs: hydroxyeicosatetraenoic acids
HPLC: high-performance liquid chromatography
HpODEs: hydroperoxyoctadecadienoic acids
LDH: lactatdehydrogenase
LTA: light transmission aggregometry
MA: maximal aggregation
MCF: maximal clot firmness
MeOH: methanol
MS: mass spectrometry
PCs: platelet concentrates
PI: pathogen inactivation
Plt: platelet
PR: pathogen reduction
rWBC: residual white blood cells
SD: standard deviation
SPE: solid-phase extraction
TA-GvHD: transfusion associated graft versus host disease
TEM: thromboelastometry
TRAP-6: thrombin receptor agonist peptide-6
TTIs: transfusion transmitted infections
yIRR: gamma irradiation

## Acknowledgements

This work was supported by the University of Vienna and by the Joint Metabolome Facility (University of Vienna, Medical University of Vienna), member of the VLSI (Vienna Life Science Instruments).

## Authorship Contributions

GCL, CG: Conceived and designed the analysis; GH, LN, AB, MH, KPW, VK, AnT: Collected the data; AlT: Contributed data or analysis tools; GH, LN, AB, CG: Performed the analysis; AB, GH, GCL, CG, WR: Wrote the paper.

