## Supplementary figures and images for "INTERCEPT pathogen reduction of platelet concentrates induces trans-arachidonic acids and affects eicosanoid formation"

### Supplementary Figure 1

#### Slide 1
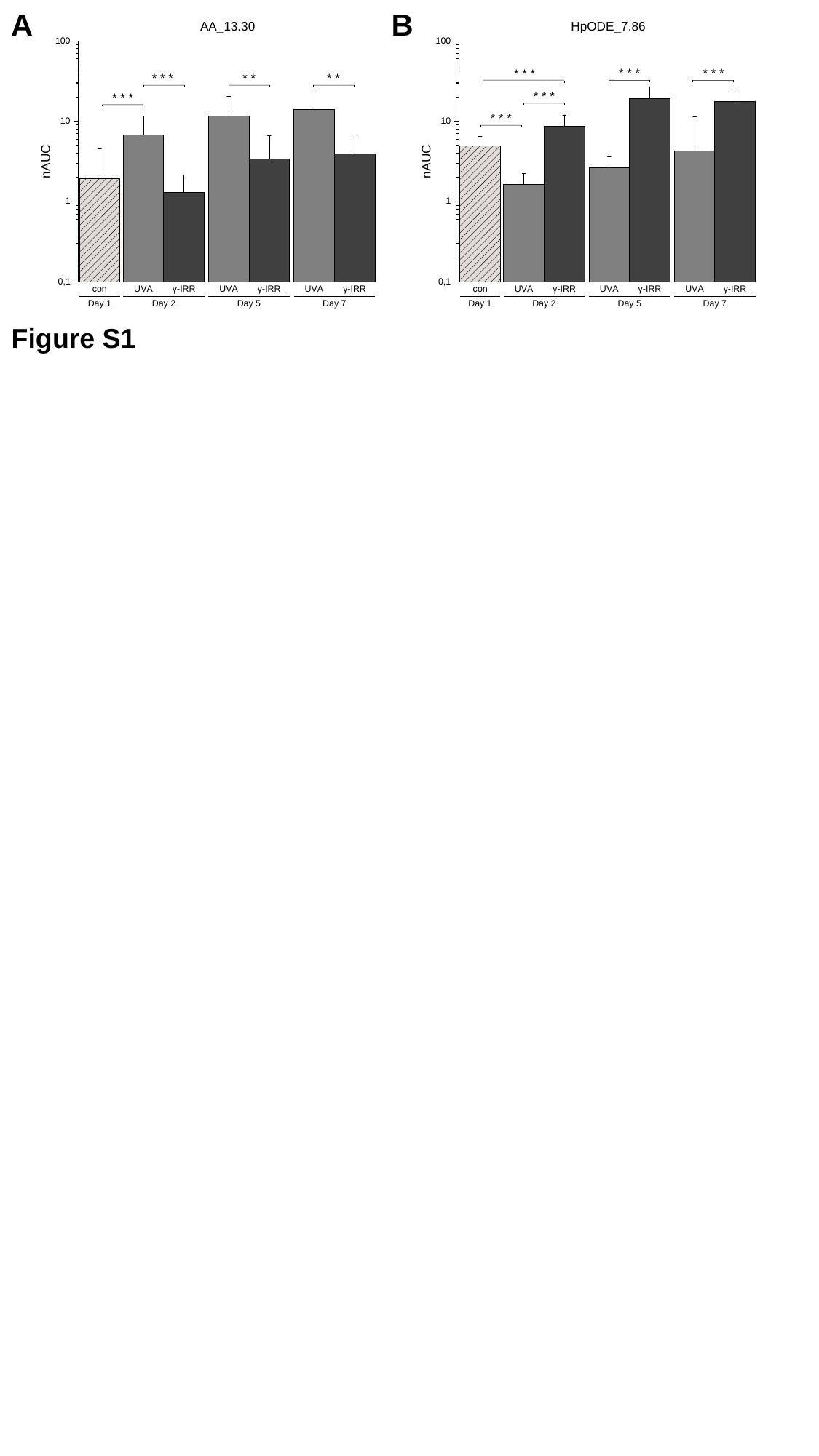

A
B
* *
* *
* * *
* * *
* * *
* * *
 * * *
* * *
* * *
Figure S1
